# Association between glycated hemoglobin A1c and automated abdominal aortic calcification: the UK Biobank Imaging Study

**DOI:** 10.64898/2026.07.02.26357193

**Authors:** Haftom N. Abraha, Abadi K. Gebre, Cassandra Smith, Lakshini Y. Herat, James Webster, Afsah Saleem, Syed Zulqarnain Gilani, Christian M. Girgis, Nicklas H. Rasmussen, William D. Leslie, John T. Schousboe, Nicholas C. Harvey, Marc Sim, Joshua R. Lewis

## Abstract

**Background:** Poor glycemic control is associated with cardiovascular disease (CVD) risk. However, it is unknown whether glycemic control is related to abdominal aortic calcification (AAC), a marker of subclinical CVD. We investigated the association between glycated hemoglobin (HbA1c) and moderate-to-high automated AAC among middle-aged to older adults from the general population.

**Methods:** We included UK Biobank Imaging Study participants free of atherosclerotic CVD at baseline. HbA1c was measured at baseline (2006-2010) and categorized as normoglycemia (<39.0 mmol/mol), prediabetes (39.0–47.9 mmol/mol), undiagnosed diabetes (HbA1c ≥48 mmol/mol) and diagnosed diabetes. Machine learning derived AAC24 (ML-AAC24) scores were estimated using a validated automated algorithm applied to dual-energy X-ray absorptiometry lateral spine images (2014-2022). The associations of HbA1c with Moderate-to-high ML-AAC24 (defined as a score ≥2) were assessed using logistic regression adjusting for cardiovascular risk factors.

**Results:** Of the included 48,912 participants (mean ± SD age 55 ± 7.6 years, 52% women), 9.7% had prediabetes (HbA1c 39.0–47.9 mmol/mol [5.7-6.4%]), 0.4% had undiagnosed diabetes and 2.7% diagnosed diabetes. Each 1-SD increase in log-transformed HbA1c was associated with higher odds of moderate-to-high ML-AAC24 (adjusted odds ratio [aOR] 1.12, 95%CI: 1.09-1.16). Amongst individuals with normal HbA1c this association was consistent but somewhat weaker for each 1-SD increase in log-transformed HbA1c (aOR 1.07, 95%CI 1.03-1.10). Compared to participants with normal HbA1c, those with prediabetes (aOR 1.19, 95%CI: 1.08-1.30) or diagnosed diabetes (1.64, 95%CI: 1.39-1.94) had higher odds of moderate-to-high ML-AAC24. These associations were consistent in stratified analyses by sex, age groups, body mass index, smoking status and total cholesterol

**Conclusions:** Linear associations between HbA1c levels and ML-AAC24 were observed in UK adults, even in those with normal HbA1c level. These findings indicate that AAC may develop early in the dysglycemic continuum, supporting earlier cardiometabolic risk assessment even amongst people with “normal” levels.

## Introduction

Poor glycemic control, including prediabetes and diabetes, affect approximately one in five adults globally ^1^. In addition to contributing to long-term microvascular complications, high glycemic levels increase the risk of cardiovascular diseases (CVD) including coronary heart disease, stroke and peripheral vascular disease ^2^. Given the rapidly increasing global prevalence of prediabetes and diabetes, understanding their relationship with CVD, the leading cause of death in this population, has become increasingly important ^3^.

Emerging evidence suggests an association between higher glycemic levels and subclinical CVD, including coronary artery calcification (CAC) ^4^ and carotid atherosclerosis ^5^ in middle-aged and older adults. Although CAC is widely used in clinical practice as a risk stratification tool for CVD, abdominal aortic calcification (AAC) often occurs before CAC ^6^, and independently predicts CV events and mortality ^7^. In the Coronary Artery Risk Development in Young Adults (CARDIA) study (n=3011, mean age ± SD 50.1 ± 3.6 years), AAC predicted incident cardiovascular events independent of CAC and even in individuals without detectable CAC ^8^. Unlike CAC, which is primarily detected by computed tomography (CT), AAC can be assessed opportunistically from low radiation lateral spine dual- energy x-ray absorptiometry (DXA) images during routine bone density testing. Importantly, CT use is limited in large-scale or screening settings due to cost, radiation exposure, and limited accessibility. Recently, AAC scoring has been automated and validated for clinical CVD prediction enabling large-scale screening for subclinical CVD ^9–11^. To date, only two small studies (<1,800 people) have investigated the relationship of HbA1c levels with AAC, and none excluded participants with prior CVD ^12,13^. Although these studies have shown the association between severe hyperglycemia and AAC, they lacked the statistical power to definitively map calcification risk within prediabetic and normal HbA1C ranges.

To better characterize the relationship between glycemic level and AAC, we investigated the association between HbA1c and machine learning AAC24 scores (ML-AAC24) in the large UK Biobank study. Specifically, we examined associations across the glycemic continuum, including normoglycemia, prediabetes, undiagnosed and diagnosed diabetes. Furthermore, we investigated *a-priori* interaction terms to identify subgroups where these associations might differ among established CVD risk factors to improve our understanding of the nature of the relationship between the glycemic continuum with ML-AAC24.

## Methods

### Study population

The UK Biobank (UKB) include 502,665 participants (age 40–69 years) recruited from 22 assessment centers across the UK ^14^. At baseline (2006-2010), participants completed health and lifestyle questionnaires (both self-completed by computer touchscreen and research nurse interview). Additionally, participants provided blood, urine, and saliva for laboratory tests ^15^. In 2014 the UK Biobank Imaging Study commenced with the aim of acquiring detailed images on 100,000 participants. Our ML-AAC24 algorithm was applied to lateral spine images (LSIs) from the first imaging visit (n = 53,611). After excluding participants with missing HbA1c data and those with prevalent atherosclerotic CVD at baseline, 48,912 participants were eligible for inclusion in the current analysis (**Supplementary Figure 1**).

### Glycemic status

Plasma HbA1c levels were measured at baseline between 2006 and 2010 and a subset of participants (n = 5,369) underwent repeat HbA1c assessments at instance 1 (2012-13) ^16^. The blood collection and sampling protocols used in this study have been previously described and validated ^17^. HbA1c was measured in mmol/mol using HPLC (Bio-Rad Variant II Turbo analyzers, Bio-Rad Laboratories, USA). HbA1c values were log-transformed to approximate a normal distribution. Additionally, we expressed HbA1c as a percentage value using the National Glycohemoglobin Standardization Program HbA1c converter ^18^.

The HbA1c categories was based on the American Diabetes Association classification thresholds: normal (<39.0 mmol/mol [<5.7%]), prediabetes (39.0-47.0 mmol/mol [5.7-6.4%]), and undiagnosed diabetes (≥ 48 mmol/mol [≥6.5%]) ^19^. Diagnosed diabetes at baseline was defined based on a validated algorithm that used baseline self-reported, primary and secondary data in UK Biobank to assign presence and type of diabetes ^14^. Details on the field IDs and ICD codes on **Supplementary Table 1.**

### Covariates

All covariates were measured at baseline (2006-2010) and include, i) sociodemographic characteristics such as age at recruitment, sex (men/women), self-reported ethnicity (White/Black/Asian/Other) and Townsend deprivation index, ii) lifestyle factors such as smoking status (never/current/previous) and physical activity using the validated short International Physical Activity Questionnaire (IPAQ) ^20^, iii) other metrics such as body mass index (BMI), systolic blood pressure (SBP), high density lipoprotein (HDL), total cholesterol, estimated glomerular filtration rate (eGFR) calculated based on serum creatinine ^21^. Self-reported antihypertensive, cholesterol lowering medications and antidiabetic medication use were also included. Detailed covariate descriptions are provided in **Supplementary Text**.

### Automated abdominal aortic calcification assessment

DXA-derived deidentified LSIs were captured using an iDXA bone density machine (GE Healthcare, Madison, WI, USA). ML-AAC24 scores were based on the Kauppila AAC24 semi-quantitative scoring system ^22^. The initial ML-AAC24 algorithm was developed using 5012 DXA-derived LSIs from four distinct cohorts (GE and Hologic machines). Image pre-processing was conducted before fine-tuning the ML-AAC24 algorithm using 497 randomly selected, de-identified images from the UK Biobank, annotated by an expert (J.T.S.). Fine-tuning was undertaken using 10-fold cross-validation of the previously developed ML-AAC24 algorithm. Each fold consisted of a training set (402 images), validation set (45 images), and test set (50 images). The ML-AAC24 scores for the remaining LSIs were then predicted by the fine-tuned algorithm. The Pearson correlation between the fine-tuned algorithm and expert scores was strong (r=0.832) as previously detailed ^10^. High agreement (intraclass correlation coefficient >0.8) was also recorded between predicted and expert scores. External validation showed that ML-AAC24 extent was strongly associated with incident CVD across multiple cohorts, including the UKB Imaging study ^9–11^. Based on established categories, the extent of ML-AAC24 was classified as: low (ML-AAC24 <2), moderate (ML-AAC24 ≥2 to <6), and high (ML-AAC24 ≥ 6) ^11,23^. Moderate-to-high was defined as ML-AAC24 score ≥2, while the presence of any ML-AAC24 was considered as ML-AAC24 score ≥1.

### Statistical analysis

Continuous variables across the HbA1c categories are presented as mean (standard deviation) and compared by ANOVA and categorical variables are presented as numbers (percentages) and compared by chi-square test. Binary logistic regression models were used to estimate odds ratio (OR) to evaluate the association between HbA1c levels (continuous and categorical) and moderate-to-high ML-AAC24. Sex stratified analysis was also undertaken. We examined the associations between HbA1c as continuous variable and the odds of moderate-to-high ML-AAC24 using restricted cubic splines, implemented via the ‘rms’ package in R. Three models of adjustment were used: (i) Model 1 adjusted for age and sex; (ii) Model 2 adjusted for Model 1 plus ethnicity, deprivation index, IPAQ and smoking; (iii) Model 3 adjusted for Model 2 plus total cholesterol, HDL cholesterol, systolic blood pressure, cholesterol lowering medication, antihypertensive use, BMI and eGFR. For each model, only people with case-complete data for all covariates were included; consequently, sample sizes differed. Statistical analysis was performed using Stata 18.0 software (Stata Corp LLC. 2023). A P-value of < 0.05 in two tailed testing were considered statistically significant.

To reduce potential treatment related confounding, sensitivity analyses were performed excluding individuals with diagnosed diabetes and those receiving antidiabetic medications. Additional analyses were conducted within each glycemic category (normal, prediabetes, undiagnosed diabetes, and diagnosed diabetes). Subgroup analyses stratified by sex (men, women), age (tertile 1, tertile 2 and tertile 3), BMI (<25, 25–30 and ≥35 kg/m²), smoking status (never, former and current), and total cholesterol (<6.2 vs ≥6.2 mmol/L).Two-way interactions were assessed by including product terms between HbA1c and each subgroup variable (age, sex, BMI, smoking status, and total cholesterol) in the models.

All analyses conducted for the primary outcome (moderate-to-high ML-AAC24) were repeated for the secondary outcome (presence of any ML-AAC24).

## Results

### Characteristics of study participants

Baseline characteristics of the participants according to glycemic status are presented in **Table 1**. Among the 48,912 participants (52.0% women) the mean ± SD age and BMI were 55.0 ± 7.6 years and 26.6 ± 4.2 kg/m^2^, respectively. Of the study cohort, 42,646 (87.2%) had normal glycemic level, 4,749 (9.7%) had prediabetes, 209 (0.4%) had undiagnosed diabetes and 1,308 (2.7%) had diagnosed diabetes. The HbA1c measurements between baseline (2006–2010) and repeat assessment (2012–2013) among participants with measurements at both time points (n = 5,369) showed moderate stability over time, (mean absolute error = 2.34 mmol/mol) **(Supplementary Table 2)**. Among the study cohort, 17,190 participants (35.1%) had any ML-AAC24, of whom 10,397 (21.3% of the total cohort) had moderate-to-high ML-AAC24.

**Table 1.** Baseline characteristics of study participants by glycemic status (based on HbA1c levels according to American Diabetes Association definition)

| Characteristic | Overall | Normal HbA1c | Prediabetes | Undiagnosed diabetes | Diagnosed diabetes | P-value |
| --- | --- | --- | --- | --- | --- | --- |
| Participants n (%) | 48,912 (100) | 42,646 (87.2) | 4,749 (9.7) | 209 (0.4) | 1,308 (2.7) |  |
| Age, years | 55.0 (7.6) | 54.5 (7.6) | 58.4 (6.6) | 56.7 (7.0) | 57.4 (7.3) | < 0.001 |
| Sex, women n (%) | 25,423 (52.0) | 22,459 (52.7) | 2,403 (50.6) | 66 (31.6) | 495 (37.8) | < 0.001 |
| Ethnicity, White n (%) | 47,259 (96.9) | 41,429 (97.5) | 4,437 (93.8) | 188 (90.0) | 1,205 (92.6) | < 0.001 |
| BMI, kg/m<sup>2</sup> | 26.6 (4.2) | 26.3 (4.0) | 28.2 (4.8) | 31.9 (5.4) | 30.2 (5.5) | < 0.001 |
| Townsend deprivation | -1.9 (2.7) | -1.9 (2.7) | -1.8 (2.8) | -1.4 (3.0) | -1.4 (2.9) | < 0.001 |
| IPAQ group n (%) |  |  |  |  |  | < 0.001 |
| Low | 7,576 (18.3) | 6,505 (18.0) | 751 (19.5) | 52 (30.2) | 268 (25.6) |  |
| Moderate | 17,347 (42.0) | 15,151 (41.8) | 1,673 (43.4) | 65 (37.8) | 458 (43.3) |  |
| High | 16,403 (39.7) | 14,589 (40.3) | 1,428 (37.1) | 55 (32.0) | 331 (31.3) |  |
| Smoking status n (%) |  |  |  |  |  | < 0.001 |
| Never | 29,549 (60.5) | 26,161 (61.5) | 2,628 (55.5) | 106 (50.7) | 654 (50.2) |  |
| Former | 16,128 (33.1) | 13,813 (32.4) | 1,667 (35.2) | 85 (40.7) | 563 (43.2) |  |
| Current | 3,123 (6.4) | 2,577 (6.1) | 442 (9.3) | 18 (8.6) | 86 (6.6) |  |
| Systolic BP, mmHg | 135.1 (17.7) | 134.8 (17.8) | 139.6 (17.4) | 144.1 (16.6) | 138.6 (15.6) | < 0.001 |
| Total cholesterol, mmol/L | 5.7 (1.1) | 5.8 (1.0) | 5.9 (1.2) | 5.9 (1.2) | 4.6 (1.1) | < 0.001 |
| HDL cholesterol, mmol/L | 1.5 (0.4) | 1.5 (0.4) | 1.4 (0.4) | 1.2 (0.3) | 1.3 (0.3) | < 0.001 |
| eGFR, ml/min/1.73 m<sup>2</sup> | 106.3 (14.2) | 106.7 (14.0) | 102.9 (14.1) | 108.6 (15.3) | 106.0 (16.0) | < 0.001 |
| Chol. lowering meds, n (%) | 5,414 (11.1) | 3,540 (8.3) | 941 (19.8) | 39 (18.7) | 894 (68.4) | < 0.001 |
| Antihypertensive use, n (%) | 7,140 (14.6) | 5,272 (12.4) | 1,125 (23.7) | 36 (17.2) | 707 (54.1) | < 0.001 |
| Any ML-AAC24, n (%) | 17,190 (35.1) | 14,143 (33.2) | 2,269 (47.8) | 85 (40.7) | 693 (53.0) | < 0.001 |
| Moderate-to-high<br>ML-AAC24, n (%) | 10,397 (21.3) | 8,255 (19.4) | 1,561 (32.9) | 65 (31.1) | 516 (39.5) | < 0.001 |

Data are presented as mean (SD) unless otherwise indicated. BMI, body mass index; IPAQ, international physical activity questionnaire; BP, blood pressure; eGFR, estimated glomerular filtration rate. Note: ML-AAC-24 presence was defined as score $\geq 1$ . ML-AAC-24 group were defined as follows: low (ML-AAC24 score = 0 and 1), moderate (ML-AAC24 score 2 to 5), and high (ML-AAC24 score $\geq 6$ ).

### Association of HbA1c with moderate-to-high ML-AAC24

The restricted cubic spline plot demonstrated a positive linear association between HbA1c (%) and moderate-to-high ML-AAC24 adjusted for covariates in model 3 (P for non-linearity = 0.490) (**Figure 1A**). The exposure response curves for HbA1c levels above 4.9%, the median HbA1c for people in the lowest quartile exhibited an upward-sloping trend. This trend was also evident after excluding individuals who were on antidiabetic medications (P for non-linearity = 0.771) (**Figure 1B**).

**Figure 1.**
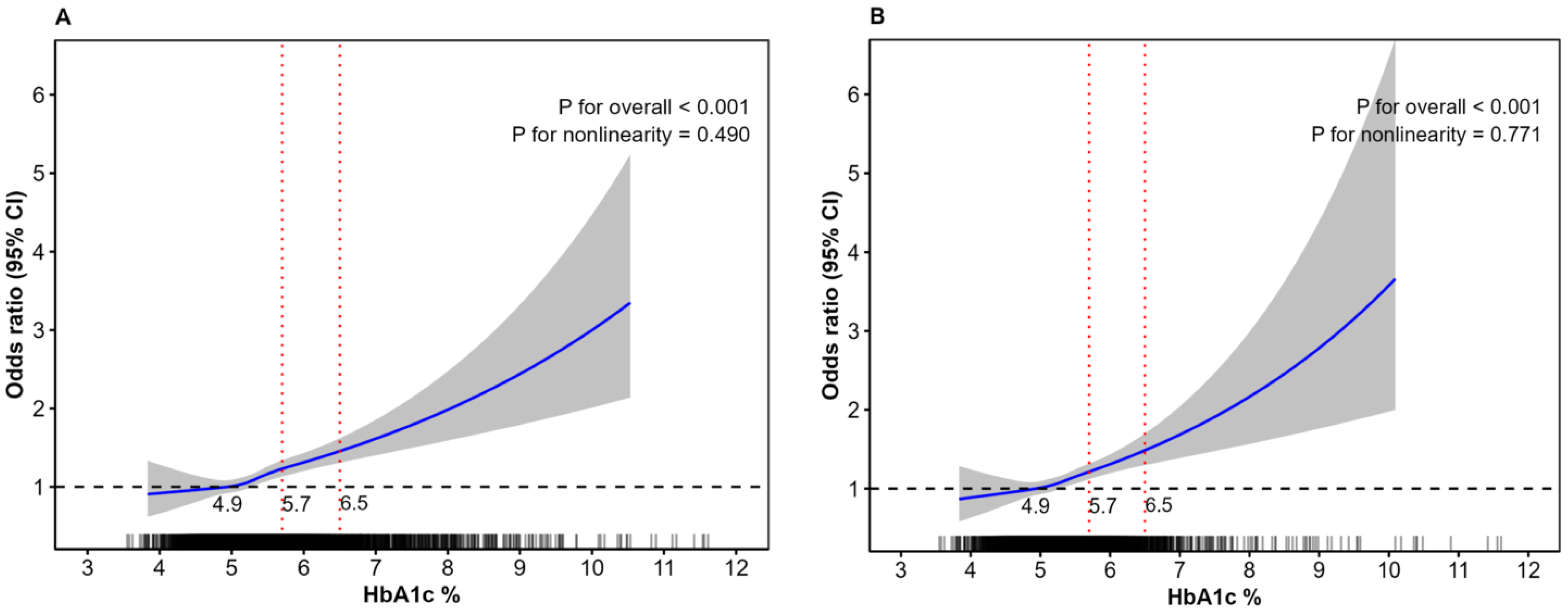
Restricted cubic splines showing the association of HbA1c % with moderate-to-high ML-AAC24 **A.** Overall (n= 32,908), **B.** Excluding people on antidiabetic medications (n= 32,344) with HbA1c= 4.9% (the median HbA1c for people in the lowest quartile) serving as the reference. The solid blue lines represent ORs, and the grey shadings represent 95% confidence regions. The rug plot along the bottom of each graph shows each observation. Multivariable-adjusted model included covariates in **Model 3** (age, sex, ethnicity, deprivation index, IPAQ, smoking, total cholesterol, HDL cholesterol, systolic blood pressure, cholesterol lowering medication, antihypertensive use, body mass index and estimated glomerular filtration rate). Vertical dashed red lines indicate the prediabetes and diabetes HbA1c cut points.

Each unit increase in SD of HbA1c (log transformed) was associated with 22% higher odds of having moderate-to-high ML-AAC24 (adjusted OR [aOR] 1.22, 95% CI: 1.19-1.26) in model 1. The odds ratio remained similar after adjustment for model 2 (aOR 1.21, 95% CI: 1.17-1.24) and slightly attenuated in model 3 (aOR 1.12, 95% CI: 1.09-1.16) (**Table 2**). The association was consistent after excluding those with diagnosed diabetes (model 3, aOR 1.09, 95% CI: 1.05-1.12) (**Supplementary Table 3**). Among individuals with normal HbA1c level (after further excluding those with HbA1c levels indicating prediabetes or undiagnosed diabetes), the association persisted (model 3, aOR 1.07, 95% CI, 1.03-1.10) (**Supplementary Table 4**).

**Table 2.**
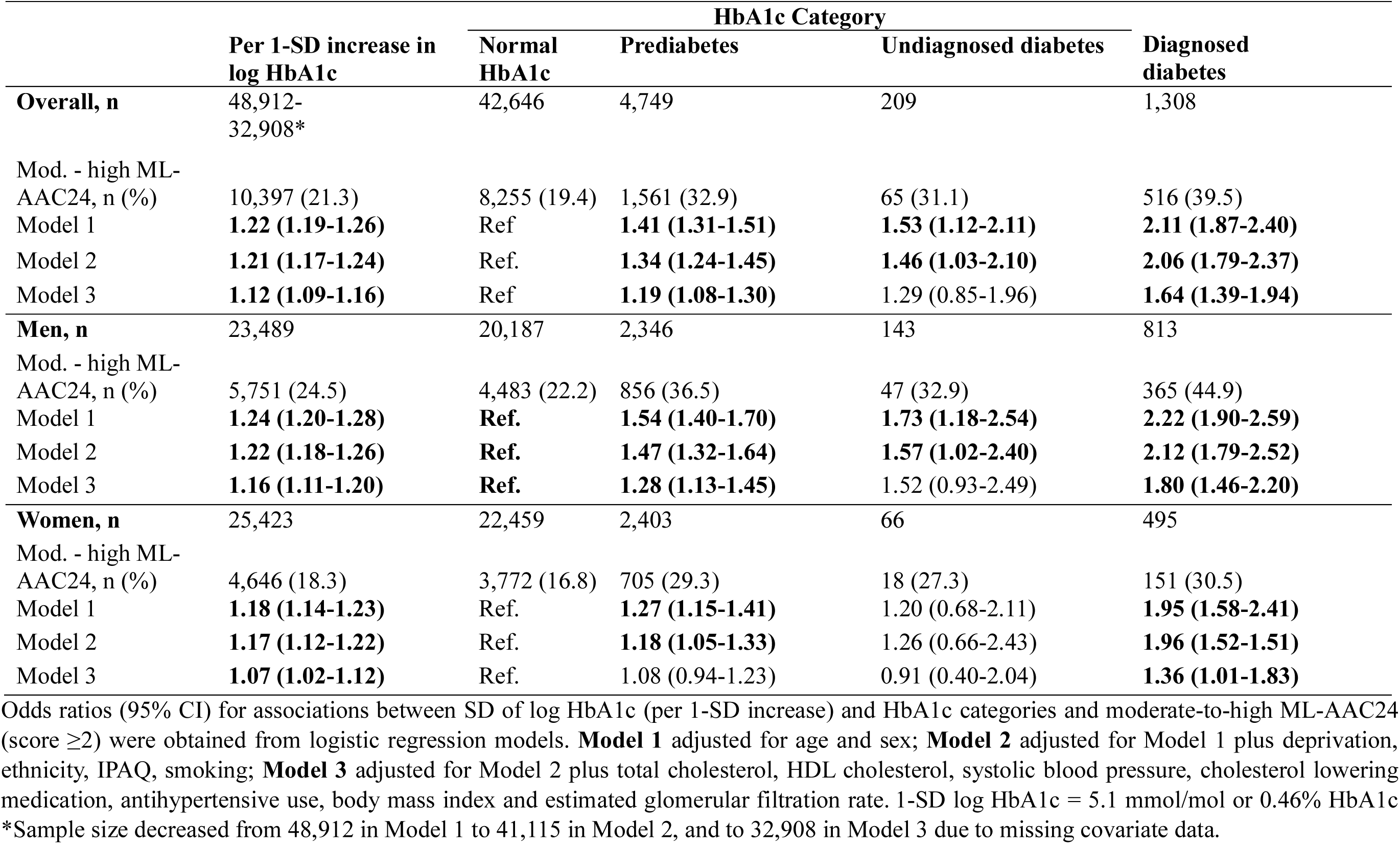
Association of HbA1c and glycemic status with odds of moderate-to-high ML-AAC24 in all study population HbA1c Category

The prevalence of moderate-to-high ML-AAC24 increased by glycemic category, from 19.4% in normoglycemia to 32.9% in prediabetes, 31.1% in undiagnosed diabetes and 39.5% in diagnosed diabetes. Logistic regression analyses revealed increased odds of having moderate-to-high ML-AAC24 across the dysglycemic categories. In age and sex adjusted analysis, compared with normoglycemia, prediabetes (aOR 1.41, 95%CI: 1.31-1.51), undiagnosed diabetes (aOR 1.53, 95%CI: 1.12-2.11), and diagnosed diabetes (aOR 2.11, 95%CI: 1.87-2.40), were associated with higher odds for moderate-to-high ML-AAC24 (**Table 2)**. The odds ratio slightly attenuated after adjustment for model 2 (prediabetes [aOR 1.34, 95%CI: 1.24-1.45], undiagnosed diabetes [aOR 1.46, 95%CI: 1.03-2.10], and diagnosed diabetes [aOR 2.06, 95%CI: 1.79-2.37]). In model 3, individuals with prediabetes and diagnosed diabetes had 19% (aOR 1.19, 95%CI: 1.08-1.30) and 64% (aOR 1.64, 95%CI: 1.39-1.94) higher odds of moderate-to-high ML-AAC24, respectively. However, statistical significance was lost in those with undiagnosed diabetes. The association between HbA1c (continuous and categorical) and moderate-to-high ML-AAC24 in model 1 and 2 remained similar when analysis was restricted for participants with no missing covariates (**Supplementary Table 5**). Furthermore, among participants with repeated HbA1c assessments, the associations of HbA1c at instance 1 (per 1-SD increase) and glycemic categories with moderate-to-high ML-AAC24 were consistent with the primary analysis (**Supplementary Table 6**).

### Subgroup analyses

We then performed the primary analyses stratified by sex, age, BMI, smoking status and total cholesterol **(Figure 2)**. Across all subgroups, each unit increase in SD of log HbA1c was associated with higher odds for moderate-to-high MLAAC24 after adjusting for covariates in model 3. The OR was relatively higher in men those in the youngest age tertile, with BMI 25-30 kg/m^2^, who were current smokers or with total cholesterol < 6.2 mmol/L although none of the interactions were statistically significant **(Figure 2)**. After exclusion of participants with diagnosed diabetes, a borderline interaction between sex and HbA1c on moderate-to-high ML-AAC24 was also observed with higher odds ratio seen in men than women (P for interaction 0.057). A significant interaction between smoking and HbA1c on moderate-to-high ML-AAC24 was also observed with highest OR in current smokers as compared to former smokers and never smokers (P for interaction 0.032) **(Supplementary Figure 2)**. Restricted cubic spline analyses stratified by smoking status demonstrated that, in current and former smokers, steeper increase in odds of moderate-to-high ML-AAC24 even at HbA1c levels below 5.7% (**Supplementary Figure 3**). Similarly, the steeper increase in OR was also observed in men compared to women participants (**Supplementary Figure 4**).

**Figure 2.**
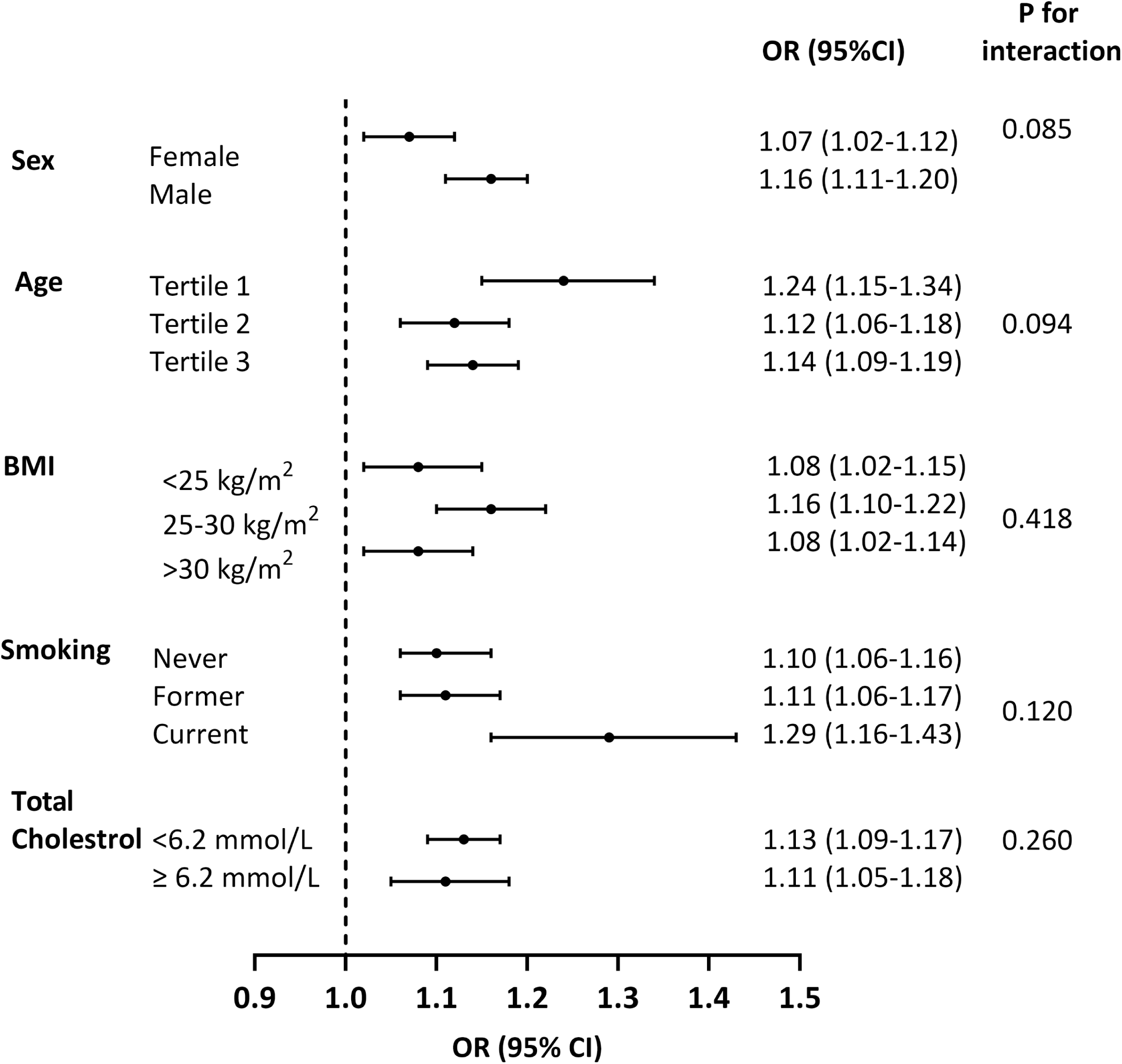
Subgroup analysis for the association of HbA1c with moderate-to-high ML- AAC24 in in participants of the UK Biobank, overall (n= 32,908). Odds ratios (95% CI) for associations between log HbA1c (per 1-SD increase) and moderate-to-high ML-AAC24 were obtained from logistic regression adjusted for covariates in **Model 3** (age, sex, deprivation, ethnicity, IPAQ, smoking, total cholesterol, HDL cholesterol, systolic blood pressure, cholesterol lowering medication, antihypertensive use, BMI and eGFR) except the corresponding stratification variable. 1-SD log HbA1c = 5.1 mmol/mol or 0.46% HbA1c

### Association of HbA1c with Any ML-AAC24

The prevalence of any ML-AAC24 increased by glycemic category, from 33.2% in normoglycemia to 47.8% in prediabetes, 40.7% in undiagnosed diabetes and 53% in diagnosed diabetes.

Restricted cubic spline plot demonstrated a positive linear association between HbA1c (%) and any ML-AAC24 adjusting for covariates in model 3 (P for non-linearity = 0.247) **(Supplementary Figure 5A)**. The exposure response curves for HbA1c levels above 4.9%, the median HbA1c for people in the lowest quartile exhibited an upward-sloping trend. This trend was also evident after excluding individuals with diagnosed diabetes (P for non-linearity = 0.374) (**Supplementary Figure 5B**).

Each one-unit increase in SD HbA1c (log transformed) was associated with 8% higher odds of having any ML-AAC24 (aOR 1.08, 95% CI: 1.06-1.11) in model 3 **(Supplementary Table 7)**. This association remained consistent after excluding participants with diagnosed diabetes (aOR 1.05, 95% CI: 1.02-1.08) in model 3 (**Supplementary Table 8**).

The categorical analyses revealed increased odds of having any ML-AAC24 across the dysglycemic categories. Compared to participants with a normal HbA1c levels, those with prediabetes and diagnosed diabetes had 19% (aOR 1.19, 95%CI: 1.09-1.30) and 60% (aOR 1.60 95%CI: 1.36-1.87) higher odds of any ML-AAC24, respectively in model 3. However, the association was attenuated for the undiagnosed diabetes **(Supplementary Table 6)**.

In subgroup analyses, each unit increase in SD HbA1c (log transformed) was associated with any ML-AAC24 across all subgroups after adjusting for covariates in model 3. Significant effect modifications by sex, BMI and smoking status were observed. The associations were more pronounced in men, those with BMI 25-30 kg/m^2^, and who were current smokers (**Supplementary Figure 6**).

## Discussion

The present analysis represents the largest and most well-characterized study to date assessing the relationship between various levels of glycemia and automatically scored AAC among community-dwelling middle aged and older adults. The key finding of the study was that higher levels of HbA1c were associated with greater odds of moderate-to-high ML-AAC24 independent of established CVD and demographic risk factors. This relationship between HbA1c and increased odds of moderate-to-high Ml-AAC24 was observed even within individuals currently considered to have “normal” glycemic levels (HbA1c <5.7%). The direction of association in all subgroups was consistent with those observed in the overall population. Collectively, robust linear associations between HbA1c levels and the extent of ML-AAC24 were observed in middle-aged and older UK adults, supporting the existing evidence of a nexus between elevated glycemic levels and subclinical CVD. These results indicate that vascular calcification may develop earlier in the dysglycemic continuum than previously thought, supporting early cardiometabolic risk assessment even amongst people with HbA1c below 5.7%.

Previous studies have shown a higher risk of AAC in those with diabetes compared to without diabetes ^12,13,24^, but little is known about the prevalence of AAC across the different stages of dysglycemia, especially in middle-aged to older adult populations. In this analysis, the prevalence of moderate-to-high ML-AAC24 was higher in those with prediabetes (32.9%) as compared with those with normoglycemia (19.4%). Notably, this prevalence was more than twice as high in individuals with diagnosed diabetes, particularly among men, in whom the prevalence was 44.9% compared to 22.2% in those with normoglycemia.

A previous study from the NHANES 2013-2014 cycle of 1,799 men and women reported that each unit increase in HbA1c% was associated with higher odds of manually-scored severe AAC (AAC-24 score > 6) (aOR 1.63, 95% CI: 1.29–2.06). However, association was not observed after excluding participants with diabetes ^13^. Additionally, two other post hoc analyses from the NHANES showed that fasting blood glucose was positively associated with AAC score (β= 0.53 95% CI: 0.28-0.78) and AAC severity (OR per SD increase 1.25, 95% CI: 1.02-1.53) ^25,26^. Furthermore, In the Jackson Heart study, a 1- SD increase in HbA1c (1.7%) was associated with higher odds of AAC presence measured by non-contrast computed tomography (prevalence ratio 1.04, 95% CI: 1.02–1.07) and diabetes but not prediabetes was associated with presence of AAC. However, all of these aforementioned studies did not exclude individuals with prior CVD and did not report whether the association persisted after excluding individuals with diabetes ^12^. After overcoming these limitations, our findings indicate a robust association across the full glycemic spectrum. Each 1-SD increase in HbA1c (log-transformed) was associated with 12% higher odds of having moderate-to-high ML-AAC24 (aOR 1.12, 95%CI: 1.09-1.16). This association remained significant even after excluding individuals with diagnosed diabetes (aOR 1.09, 95%CI: 1.05-1.12). Notably, the association persisted among individuals with HbA1c levels within the normal range (i.e. HbA1c <5.7%), where each 1-SD increase in HbA1c (log-transformed) was associated with 7% increase in the odds of moderate-to-high ML-AAC24 (aOR 1.07, 95% CI: 1.03-1.10). These findings suggest that glycemic exposure, even below the diagnostic threshold for prediabetes and diabetes, may play an important role in vascular calcification and cardiovascular risk. In line with the current findings, a prospective cohort study (median follow-up 14 years) has shown that the risk of developing coronary heart diseases and stroke may begin at HbA1c levels (5.5 to 6.4%), this suggests that CVD disease is developing below and potentially well below the diagnostic threshold for diabetes ^27^.

HbA1c is widely recognized as a reliable marker for assessing average plasma glucose concentrations over a period of 2 to 3 months and a value of 6.5% or greater indicates a T2DM diagnosis ^28^. This diagnostic threshold, however, is based on the association of plasma glucose levels and microvascular complications of diabetes such as retinopathy, rather than macrovascular complications such as stroke and acute coronary syndromes ^29^. Many studies have shown that prediabetic levels of HbA1c are associated with an increased risk of subclinical ^13,30^ and clinical CVD ^2,27,31^. Recent meta-analysis of prospective cohort studies and clinical trials of adults involving nearly 10 million participants indicate that individuals with prediabetes, defined by elevated fasting blood glucose or HbA1c levels, are at a greater risk of developing clinical CVD ^2^. A previous study on UK Biobank participants also reported that, compared to individuals with HbA1c <6%, those with prediabetes and undiagnosed diabetes had a significantly higher risk of developing clinical CVD ^31^. Our study has confirmed HbA1c is associated with AAC independent of CVD risk factors. Due to its reliability, extensive availability, and cost effectiveness, HbA1c measurements can serve as useful marker for assessing subclinical CVD in individuals without a diagnosis of diabetes. Given that HbA1c levels serve as the “gold standard” for evaluating glycemic control ^19^, our findings underscore the potential added value of glycemic level to early identify individuals at higher risk of subclinical and clinical CVD.

Multiple potential mechanisms may contribute to the association between elevated glycemic levels and AAC. Hyperglycemia contributes to the development of intimal calcification by promoting endothelial dysfunction, raising oxidative stress, and triggering proatherogenic pathways ^32^. It also plays and active role in the trans-differentiating vascular smooth muscle cells into an osteoblast-like phenotype, enhancing of extracellular matrix calcification and upregulation of bone matrix proteins leading to the initiation and progression of medial calcification ^33^.

In a subgroup analysis excluding individuals with diagnosed diabetes, smoking status interacts with HbA1c level for the likelihood of ML-AAC24 (P-for interaction= 0.032). The association between elevated HbA1c and ML-AAC24 was stronger among current smokers compared to former smokers or non-smokers, suggesting potential toxic damage from cigarette smoke markedly elevates risk of vascular calcification. This finding is consistent with prior prospective studies which showed the interaction between smoking and diabetes on CVD events ^34^. As shown in the stratified restricted cubic spline, the higher risk of moderate-to-high ML-AAC24 in current smokers with HbA1c levels <5.7% suggests that, smoking with slightly elevated glycemia within normal levels may amplify the vascular calcification risk, potentially by promoting further inflammation, endothelial dysfunction, and increased production of reactive oxygen species ^35^. In addition to creating pro-calcific environment, smoking overwhelms and functionally impairs the endogenous vascular calcification inhibitors such as matrix Gla protein and osteocalcin (bone Gla protein) ^36^. Experimental studies are needed to shed light on potential mechanisms responsible for the interplay between smoking and dysglycemia on AAC risk.

This study adopted a large sample size from middle-aged to older adults without overt CVDs and accounted for established CVD risk factors and medications (antidiabetic, cholesterol lowering, antihypertensive) to minimize confounding. The previous studies had small sample sizes as they were based on manual AAC scoring, but the automated AAC scoring enabled application in a large cohort study. We utilized a state-of-the-art machine learning algorithm for assessing AAC that was validated with a human expert and has been shown to strongly and robustly predict incident clinical atherosclerotic CVD outcomes in the UK biobank ^9^. Furthermore, we investigated the association across the glycemic continuum and in pre-specified subgroups to look for robustness and consistency. However, there are some limitations that should be noted. Firstly, the UK Biobank comprises a predominantly White population, which may limit the generalizability of our findings to other ethnic groups. Secondly, causality cannot be determined due to the observational nature of the study. Thirdly, there was a time gap between the baseline HbA1c measurement (2006–2010) and AAC assessment (2014-2022). However, in the 11% of participants with repeat HbA1c in 2012–2013, values were stable, supporting baseline HbA1c as a reasonable proxy for long-term glycemic exposure. Finally, despite adjustments for known risk factors using multivariate models there may have been residual confounding from both unmeasured and measured variables. For example, the baseline period largely predates widespread use of GLP-1 receptor agonists and SGLT2 inhibitors. In contrast, imaging occurred during an era of more intensive cardiometabolic prevention, including broader use of statins and GLP-1 receptor agonists for cardiovascular risk reduction. This might have raised the possibility of unmeasured treatment-related confounding.

In conclusion, robust associations between HbA1c and ML-AAC24 were observed in middle-aged and older UK adults across the full glycemic spectrum, further strengthening the evidence that dysglycemia is associated with the development and progression of CVD. This study confirms a strong association between poorer glycemic control and vascular calcification, which may emerge earlier in the dysglycemic spectrum than previously recognized. Future longitudinal and interventional studies are warranted to further investigate this association with subclinical CVD in other vascular beds and whether HbA1c lowering may reduce the development and progression of subclinical CVD and ultimately clinical CVD risk.

## Data Availability

The data that support the findings of this study are available from UK Biobank but restrictions apply to the availability of these data, which were used under license for the current study, and so are not publicly available. Data are, however, available from the principal investigator (JRL) upon reasonable request and with permission from UK Biobank.

## Acknowledgements

The authors thank the participants of the UK Biobank

## Authors contributions

H.A. wrote the first draft of the manuscript. A.G., C.S., L.H., and J.L., were involved in the conception, design, and conduct of the study and the analysis and interpretation of the results. M.S. was involved in the conception and design of the study. H.N., and A.G. conducted statistical analyses. All authors edited, reviewed, and approved the final version of the manuscript.

## Ethics

Ethics approval for the UK Biobank was obtained from the Northwest Multicenter Research Ethics Committee UK (REC reference: 11/NW/03/820). Written informed consent for data collection and record linkage was obtained from all UKB participants. The study was conducted in accordance with the principles of the Declaration of Helsinki.

